# COVID-19 Pandemic Preparedness in a United Kingdom Tertiary and Quaternary Children’s Hospital: Tales of the Unexpected

**DOI:** 10.1101/2020.08.20.20178541

**Authors:** Nele Alders, Justin Penner, Karlie Grant, Charlotte Patterson, Jane Hassell, Nathalie MacDermott, Sian Pincott, Alasdair Bamford, Pascale du Pré, Mae Johnson, Karyn Moshal

## Abstract

**Background:** The paucity of data describing SARS-CoV-2 in the paediatric population necessitated a broad-arching approach to pandemic planning, with preparations put in place to manage a heterogeneous cohort. We describe a diverse group of SARS-CoV-2 positive paediatric patients treated at a large tertiary/quaternary children’s hospital in the United Kingdom and the adaptive coping strategies required.

**Methods:** All paediatric patients with positive RT-PCR on a respiratory sample and/or serology for SARS-CoV-2 up to 19^th^ May 2020 were included.

**Results:** 57 children met the inclusion criteria. 70% were of non-Caucasian ethnicity with a median age of 9.3 years (IQR 5.16–13.48). Four distinct groups were identified: paediatric inflammatory multisystem syndrome temporally associated with SARS-CoV-2 (PIMS-TS) (54%), primary respiratory (18%), incidental (7%), and non-specific febrile illnesses with or without extra-pulmonary organ dysfunction (21%). These groups presented in distinct chronological blocks as the pandemic unfolded.

**Discussion:** The diverse range of presentations of SARS-CoV-2 infection in this population exemplified the importance of preparedness for the unknown in the midst of a novel infectious pandemic. Descriptions of paediatric patients during the initial phase of the pandemic from other parts of the globe and extrapolation from adult data did not serve as an accurate representation of paediatric COVID-19 in our centre. An adaptive, multidisciplinary approach was paramount. Expanded laboratory testing and incorporation of technology platforms to facilitate remote collaboration in response to strict infection control precautions were both indispensable. Lessons learned during the preparation process will be essential in planning for a potential second wave of SARS-CoV-2.

## Introduction

SARS-CoV-2, a novel zoonotic coronavirus causing severe respiratory symptoms in adults (coronavirus disease 2019 [COVID-19]) was first identified in China in December 2019. The first cases of COVID-19 in the United Kingdom (UK) were identified on 29^th^ January 2020 with the World Health Organisation (WHO) subsequently declaring COVID-19 a pandemic on 11^th^ March 2020. Data from England suggests that 1.7% and 0.8% of COVID-19 cases affected people under twenty and ten years of age respectively, with increasing rates of positive cases in children first noted in March 2020 (1).

The relative paucity of data describing SARS-CoV-2 in the paediatric population mandates a broad-arching approach to pandemic planning with preparations put in place to manage a heterogeneous population of patients presenting with a range of single and multi-organ pathology of varying severity. We describe a diverse group of SARS-CoV-2 infected paediatric patients treated at Great Ormond Street Hospital, a tertiary and quaternary paediatrics hospital in London, UK with 383 inpatient beds and approximately 50 specialties, whom exemplified the importance of preparedness for the paediatric COVID-19 unknown. We further illustrate four distinct temporal waves of SARS-CoV-2 clinical phenotypes at our centre beginning with our first case 25^th^ March 2020.

## Methods

All patients aged ≤ 18 years with positive respiratory or nasal SARS-CoV-2 RT-PCR and/or serum IgG (Epitope Diagnostics Inc.^TM^) up to 19^th^ May 2020. This time interval was chosen as it corresponds to the first two months of local paediatric cases when preparation measures remained in flux. It also represents the onset of the local epidemic when community seroprevalence remained low, thus, both diagnostic methods are likely to represent recent infections. A search of electronic medical records for clinical, laboratory, and radiographic data was performed. False-positive results due to lab contamination were excluded.

Anonymised data was collected and stored in a secure Excel^®^ database. The project was registered with the local research department (approval #2857). Real-time laboratory data was collected via the microbiology tracking system of the electronic patient records.

## Results

2,194 SARS-CoV-2 RT-PCR tests had been undertaken on 933 patients. Antibody tests were also introduced on-site assessing IgG response to SARS-CoV-2 nucleocapsid (sensitivity 92%, specificity 96%). 126 patient antibody tests had been completed.

Of 65 paediatric patients with positive samples (n = 28 RT-PCR, n = 27 serology, n = 10 both), 57 patients were included in the final analysis. Three patients were excluded for unconfirmed positive nasopharyngeal aspirates (NPA) reported prior to hospital transfer with negative admission screening. One serology-positive infant with negative RT-PCR was excluded because of uncertainty in the significance of the result. The mother had symptoms consistent with COVID-19 late in the third trimester, therefore passive maternal antibody transfer was suspected. Three patients were excluded due to lack of data as they did not require hospital admission

### (i) Clinical & Laboratory Characteristics

The cohort characteristics are presented in *Table 1*. The median age was 9.3 years (IQR 5.16–13.48) and 70% were of non-Caucasian or mixed ethnicity. Four distinct clinical groups were identified: paediatric inflammatory multisystem syndrome temporally associated with SARSCoV-2 (PIMS-TS) (54%), primary respiratory (18%), incidental (7%), and non-specific febrile/viral illness with or without single organ dysfunction (21%). These groups presented in distinct chronological blocks (*Figure 1*) with respiratory and other febrile illnesses predominating in the first three weeks after the first positive case was admitted. This was followed almost exclusively by PIMS-TS cases in the latter third of the study period. Compared to those with a primary respiratory phenotype, PIMS-TS patients were generally older (median 10.1 [8.7–13.9] vs. 3.4 years [0.1–8.2]) and of non-Caucasian ethnicity (n = 26 [84%] vs n = 5 [50%]). 61% had no known contacts with COVID-19. All incidental cases had household exposure whereas household exposure was less common in the remainder of the groups (29–33%). The most commonly demonstrated symptoms (prior to or during admission) comprised of: fever (94%), vomiting(72%), abdominal pain (64%), diarrhoea (53%), and rash (49%). Upper and lower respiratory tract symptoms were present in 30% and 40% of patients respectively, predominantly amongst primary respiratory phenotypes, though still present in all symptomatic groups. Systemic inflammatory signs often seen in conditions such as Kawasaki’s disease were common, as were central nervous system symptoms (55% of symptomatic patients ≥ 1 neurological sign/symptom) such as: headache (38%), encephalopathy (32%), weakness (25%), and meningism (9%). Abdominal and neurologic symptoms were mostly seen in PIMS-TS where conjunctivitis and rash were almost exclusive to this group. 43% were overweight (weight-forage > 85%) or obese (weight-for-age > 95%) and 9% were severely obese (weight-for-age > 99^th^ percentile). Obesity was equally common amongst PIMS-TS and respiratory phenotypes and largely restricted to these two groups. For newly admitted patients with community acquisition (n = 50), median length of stay was 9 days (IQR 6–15.5). This was longest for respiratory presentations (16.5 days [9.5–18.8]) compared to other symptomatic groups (9 [7–15] PIMS-TS, 4.5 days [2.3–10.8] other).

**Table 1:**
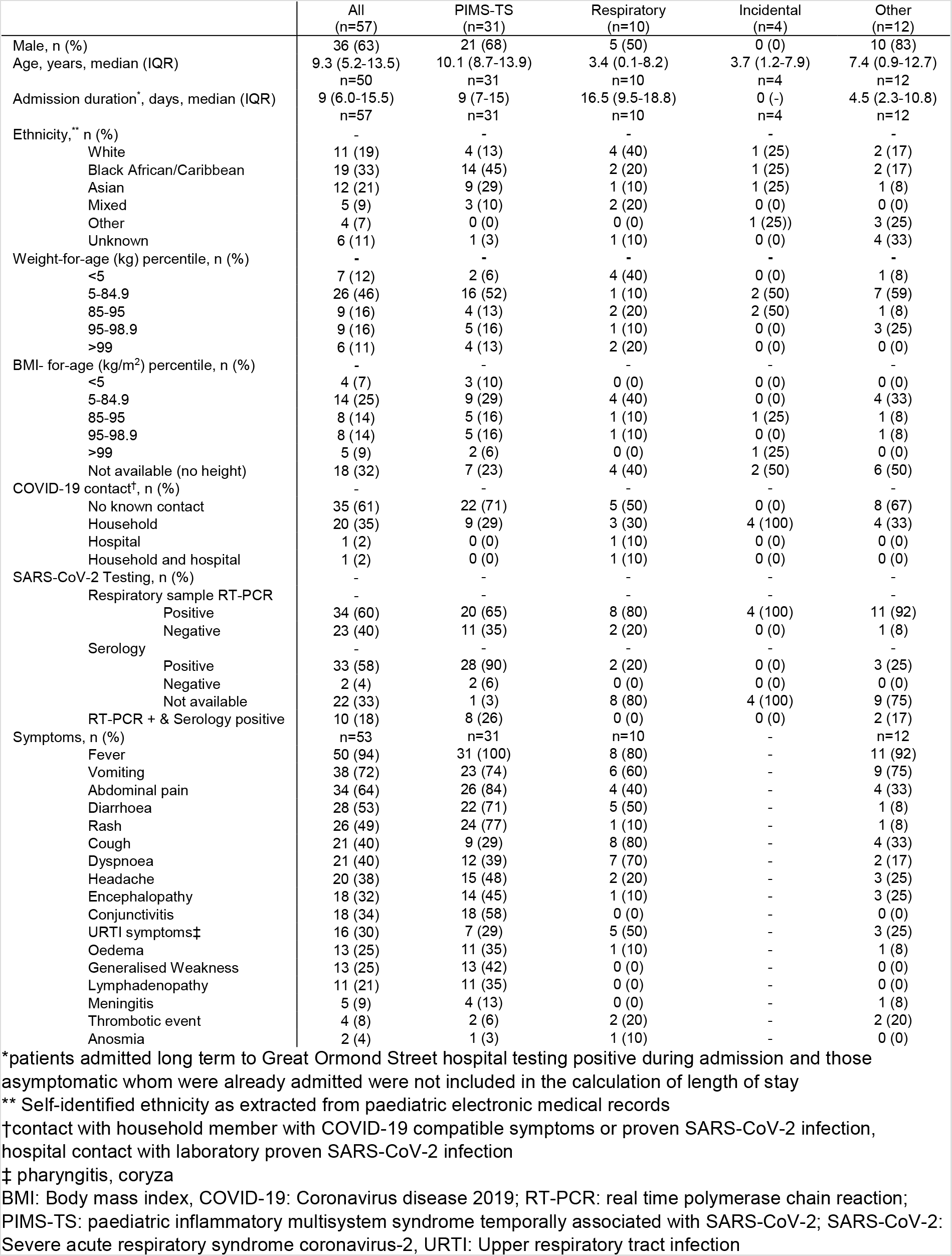
Cohort demographics

**Figure 1:**
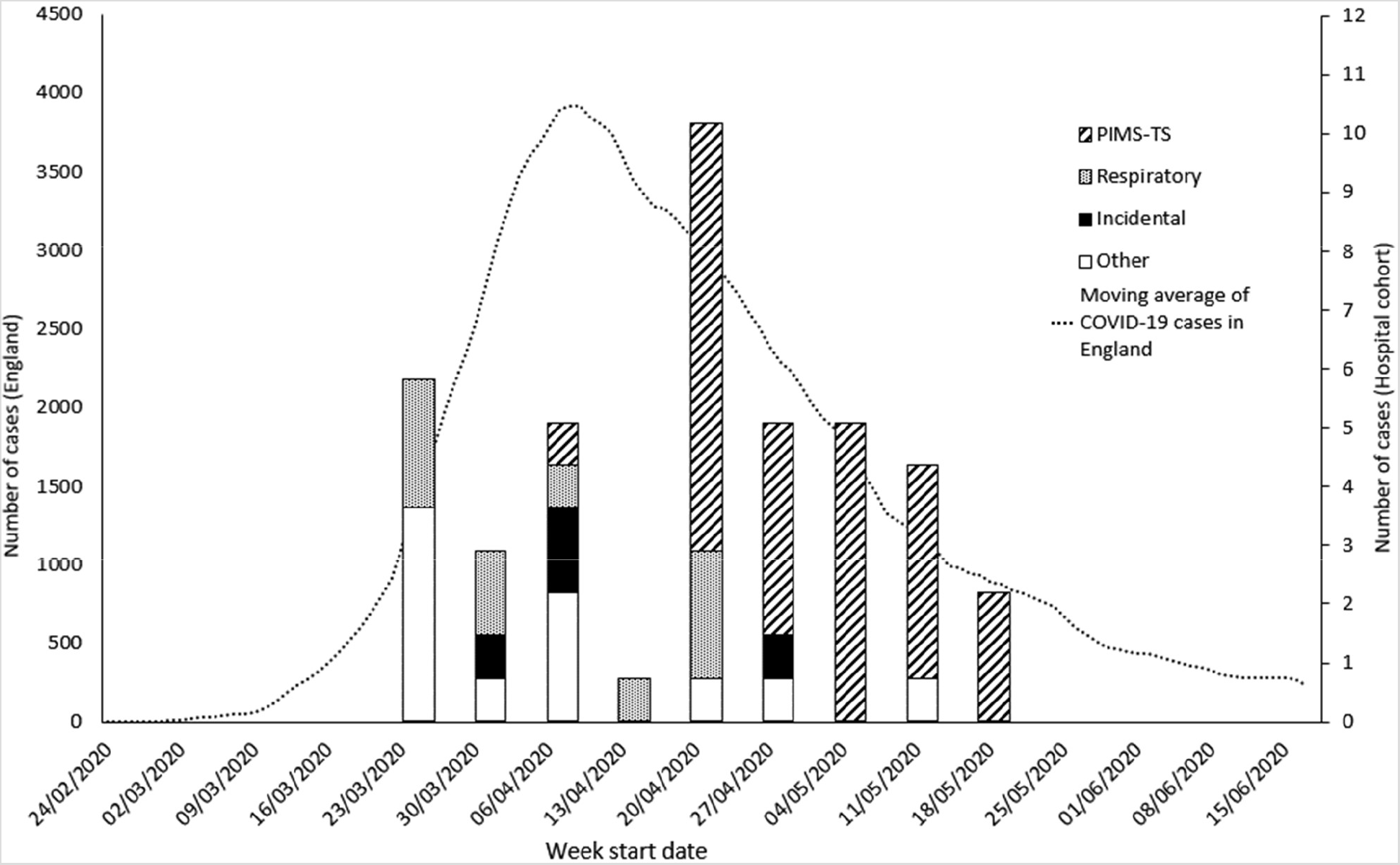
Local phenotypic presentations of paediatric SARS-CoV-2 cases over time compared to daily cases in England^1^

Laboratory and radiographic results are presented in *Table 2*. Markers of inflammation in symptomatic patients were high with a median maximum CRP of 250 mg/L (IQR), equally high amongst respiratory (258.5 [55.5–281] and PIMS-TS phenotypes (290 [213–324.5]). Hypoalbuminaemia alongside elevated fibrinogen, ferritin, and LDH were generally found in both PIMS-TS and respiratory groups with D-dimers highest for PIMS-TS (PIMS-TS: 4981 [2664–6288]; Respiratory: 1401 [1160–1332]). Lymphopaenia, thrombocytosis, and thrombocytopenia were frequent amongst all groups. Elevation of liver enzymes was apparent, especially in the latter/convalescent phase of PIMS-TS.

Two patients were found to have detectable SARS-CoV-2 RT-PCR in stool. SARS-CoV-2 was not found in urine or blood on RT-PCR though this was not routinely requested. In addition to standard bacterial cultures (blood, urine), concurrent viral infections were routinely screened for in all patients. Anti-streptolysin-O titres and specific bacterial blood PCRs were performed (meningococcus, staphylococcus aureus, group A streptococcus, and pneumococcus) in those presenting with shock. Epstein-Barr virus was the most common co-infection (n = 8).

**Table 2:** Summary of investigation results

|  | All | PIMS-TS | Respiratory | Incidental | Other | Reference range |
| --- | --- | --- | --- | --- | --- | --- |
| Blood investigations [Median, (IQR)] | n=54* | n=31* | n=10* | n=3* | n=10* | - |
| Lowest | - | - | - | - | - | - |
| Total WCC (x10<sup>9</sup>/l) | 7.0 (4.6-9.9) | 7.8 (5.1-10.2) | 7.4 (3.5-10) | 2.0 (1-4) | 6.4 (4-9.9) | 4.5-13.5 |
| Lymphocyte count (x10<sup>9</sup>/l) | 1.0 (0.5-1.6) | 0.8 (0.4-1.1) [n=30] | 1.3 (1-2.5) | 0.3 (0.1-1.2) | 1.8 (1-3.3) | 1.5-7 |
| Platelet count (x10<sup>9</sup>/l) | 164 (98.8-248.3) | 161(95-227) | 152 (111.5-239.8) | 4.0 (2.5-66.5) | 239 (182-256.8) | 150-450 |
| Albumin (g/l) | 25 (23-30.3) [n=44] | 25 (23-27.8) [n=30] | 27 (21.3-29) [n=7] | 36.5 (33.8-39.3) [n=2] | 34 (32-36) [n=5] | 37-56 |
| Highest | - | - | - | - | - | - |
| Total WCC (x10<sup>9</sup>/l) | 16.7 (12.8-24) | 21.1 (15.3-27.1) [n=30] | 15.0 (13.9-18) | 15.5 (10.8-29.8) [n=3] | 9.7 (5.4-15.6) | 4.5-13.5 |
| Neutrophil count (x10<sup>9</sup>/l) | 11.9 (6.1-16.4) [n=53] | 14.9 (11.4-25.2) | 9.2 (6.4-11.6) | 15.1 (9.1-29) | 4.4 (2.9-8.8) | 1.5-8 |
| Platelet count (x10<sup>9</sup>/l) | 457.0 (299.3-656.5) | 515.0 (397.5-684) | 533 (263.8-417.5) | 266 (197.5-282) | 319.5 (263.8-417.5) | 150-450 |
| Fibrinogen (g/l) | 5.7 (4.4-6.7) [n=49] | 6.1 (4.9-7) [n=30] | 5.5 (4.1-6.4) | 3.7 (3.5-3.9) [n=2] | 3.9 (4.1-5) [n=7] | 1.7-4 |
| D-dimer (ug/l) | 3299.0 (1160-5852) [n=45] | 4981 (2664-6288) [n=29] | 1401 (1160-2332) [n=9] | 212.0 (-) [n=1] | 375.5 (282.8-542.5) [n=6] | 0-312 |
| Ferritin (ug/l) | 818.5 (404-1585.5) [n=42] | 990 (526-2096) [n=29] | 1452 (729.5-1557) [n=7] | - | 82.7 (30.1-614.2) [n=6] | 21-92 |
| LDH (U/l) | 1016 (797-1272 [n=45] | 1119 (893-1303) | 986 (839.5-1462.5) [n=7] | 720 (646-794) [n=2] | 629 (423-695) [n=5] | 380-770 |
| CRP (mg/l) | 250 (54-301) [n=53] | 290 (213-324.5) | 258.5 (55.5-281) | 57 (28.5-65.5) | 18 (0-33) [n=9] | <5.0 |
| ALT (U/l) | 68.5 (42.5-118.5) [n=50] | 88 (48-133.5) | 71 (44-75) [n=9] | 147 (82-382.5) | 24 (19.5-43) [n=7] | 10-55 |
| AST (U/l) | 73 (46-164) [n=17] | 70 (58-84.5) [n=7] | 74.5 (41-167) [n=8] | 1500 [n=1] | 89 [n=1] | 10-40 |
| Other | - | - | - | - | - | - |
| Vitamin D (nmol/L) | 27 (16-56) [n=25] | 26 (14.8-50) [n=20] | 141 (-) [n=1] | 55.5 (42.8-68.3) [n=2] | 20.5 (13.8-27.3) [n=2] | 50-200 |
| X-ray Chest (n, %) | n=47 | n=30 | n=9 | n=2 | n=6 | - |
| Normal | 20 (43) | 12 (40) | 1 (11) | 2 (100) | 5 (83) | - |
| COVID-19 infiltrates** | 16 (34) | 7 (23) | 8 (89) | 0 (0) | 1 (17) | - |
| Pulmonary oedema | 7 (15) | 7 (23) | 0 (0) | 0 (0) | 0 (0) | - |
| Cardiomegaly | 6 (13) | 6 (20) | 0 (0) | 0 (0) | 0 (0) | - |
| Computed tomography (n, %) | - | - | - | - | - | - |
| Chest | n=6 | n=4 | n=1 | - | n=1 | - |
| Normal | 1 (17) | 1 (25) | 0 (0) | - | 0 (0) | - |
| COVID-19 infiltrates† | 5 (83) | 3 (75) | 1 (100) | - | 1 (100) | - |
| Abdomen | n=6 | n=6 | - | - | - | - |
| Normal | 2 (33) | 2 (33) | - | - | - | - |
| Ileocolitis | 4 (67) | 4 (67) | - | - | - | - |
| Ultrasound Abdomen (n, %) | n=28 | n†=23 | n=1 | n=1 | n=3 | - |
| Normal | 11 (39) | 7 (30) | 1 (100) | 1 (100) | 2 (67) | - |
| Ascites | 12 (43) | 12 (52) | 0 (0) | 0 (0) | 0 (0) | - |
| Ileocolitis | 3 (11) | 2 (9) | 0 (0) | 0 (0) | 1 (33) | - |
| Mesenteric adenitis | 3 (11) | 3 (13) | 0 (0) | 0 (0) | 0 (0) | - |
| Transthoracic Echocardiography (n, %) | n=36 | n=30 | n=4 | - | n=2 | - |
| Normal | 25 (69) | 20 (67) | 3 (75) | - | 2 (100) | - |
| Valve regurgitation | 10 (28) | 9 (30) | 1 (25) | - | 0 (0) | - |
| Abnormal Z'-scores >2 | 7 (19) | 6 (20) | 1 (25) | - | 0 (0) | - |
| Myocardial dysfunction† | 6 (17) | 6 (20) | 0 (0) | - | 0 (0) | - |
| Pericardial fluid | 4 (11) | 4 (13) | 0 (0) | - | 0 (0) | - |
| MRI Brain (n†, %) | n=14 | n=10 | n=1 | - | n=3 | - |
| Normal | 3 (21) | 1 (10) | 0 (0) | - | 2 (67) | - |
| Acute signal changes | 7 (50) | 7 (70) | 0 (0) | - | 0 (0) | - |
| Volume loss | 3 (21) | 2 (20) | 1 (100) | - | 0 (0) | - |
| Acute diagnostic changes | 2 (14) | 0 (0) | - | - | 0 (0) | - |
| EEG (n, %) | n=17 | n=12 | - | n=1 | n=4 | - |
| Normal | 2 (12) | 1 (8) | - | - | 1 (25) | - |
| Encephalopathy | 14 (82) | 11(92) | - | 1 (100) | 3 (75) | - |

\*Unless otherwise stated
\*\*COVID-19 infiltrates defined by radiology report or infiltrates consistent with previously described radiologic abnormalities seen in COVID-19 in the context of clinically consistent disease
† Patients may have more than single findings and thus classified in more than one category
ALT: alanine aminotransferase; AST: aspartate aminotransferase; COVID-19: coronavirus disease 2019; CRP: C-reactive protein; EEG: electroencephalogram; IQR: interquartile range; LDH: lactate dehydrogenase; MRI: magnetic resonance imaging; PIMS-TS: paediatric inflammatory multisystem syndrome temporally associated with SARS-CoV-2; SARS-CoV-2: severe acute respiratory syndrome coronavirus 2; WCC: white cell count

Moreover, 30 cytokine profiles (BD FACSCanto^TM^) were completed on 13 patients. Interferon gamma, IL4, IL2, and tumour necrosis factor alpha were universally < 50 pg/ml. Elevated IL10 was observed in one patient with a primary respiratory presentation [110pg/ml] and one PIMSTS (55pg/ml). Elevated IL-6 was demonstrated in six patients, four PIMS-TS (range: 59–443 pg/ml) and two primary respiratory (range: 71–218 pg/ml).

In those who underwent abdominal imaging and echocardiograms, gut inflammatory pathology on ultrasound (61%) and inflammatory heart complications (31%) were almost exclusive to the PIMS-TS group. Brain MRI (n = 14) and EEGs (n = 17) were performed more routinely in the latter part of the local epidemic when subtle neurological complications were recognised as common. Abnormal findings were seen in 79% and 82% of those conducted respectively, again almost exclusive to the PIMS-TS group. However, these tests were conducted less frequently in the early stages of the local epidemic thus leading to few MRIs and EEGs in respiratory and other presentations.

Treatment modalities are outlined in *Table 3*. Four patients were treated off-label with remdesivir via a compassionate access programme of which pK studies were conducted in three (sample analysis outstanding). All four were admitted to paediatric/cardiac intensive care (PICU/CICU) with primary respiratory disease meeting the 2015 PALICC criteria for severe paediatric acute respiratory distress syndrome (pARDS) (2). Twenty-nine (52%) were treated with immunoglobulins all of which were in the PIMS-TS group apart from one ‘other group’ treated for acute demyelinated encephalomyelitis (ADEM). Twenty-nine (52%) were treated with steroids, all were of PIMS-TS phenotype apart from two in the respiratory group where steroids were used for blood pressure support and the patient treated for ADEM. Anakinra(11%) was used off-label exclusively in PIMS-TS. Prophylactic low molecular weight heparin (LMWH) was prescribed in 56% of patients, more routinely in the later part of our local paediatric epidemic when clotting complications (4/57) were increasingly recognised.

**Table 3:** Summary of utilised management interventions

|  | All<br>(n=57) | PIMS-TS<br>(n= 31) | Respiratory<br>(n=10) | Other<br>(n=12) |
| --- | --- | --- | --- | --- |
| Methylprednisolone* n (%) | 29 (51) | 26 (84) | 2 (20) | 1 (8) |
| High dose | 22 (76) | 21 (68) | 0 (0) | 1 (8) |
| Low dose | 7 (24) | 5 (16) | 2 (20) | 0 (0) |
| Immunoglobulins | 29 (51) | 28 (90) | 0 (0) | 1 (8) |
| Anakinra | 6 (11) | 6 (19) | 0 (0) | 0 (0) |
| Remdesivir | 4 (7) | 0 (0) | 4 (40) | 0 (0) |
| Antibiotics | 49 (86) | 27 (87) | 9 (90) | 8 (67) |
| Aspirin† | 20 (35) | 20 (65) | 0 (0) | 0 (0) |
| High dose | 4 (7) | 4 (13) | 0 (0) | 0 (0) |
| Low dose | 16 (28) | 16 (52) | 0 (0) | 0 (0) |
| Low molecular weight<br>heparin** | 32 (56) | 25 (81) | 6 (60) | 1 (8) |
| Prophylactic dose | 28 (49) | 23 (74) | 4 (40) | 1 (8) |
| Treatment dose | 4 (7) | 2 (6) | 2 (20) | 0 (0) |
| Supplemental Oxygen | 29 (51) | 18 (58) | 8 (80) | 3 (25) |
| Mechanical ventilation | 21 (37) | 14 (45) | 4 (40) | 3 (25) |
| Inotropes | 23 (40) | 20 (65) | 2 (20) | 1 (8) |
| ECMO | 1 (2) | 1 (3) | 0 (0) | 0 (0) |

Incidental cases (n=4) did not receive any management interventions
\*High dose is defined as 10-30mg/kg/day, max 1g; low dose as 0.4mg/kg/day- 4mg/kg/day
\*\*High dose is defined as 30-50mg/kg/day, Low dose is defined as 3-5 mg/kg/day
†Treatment dose defined as dalteparin 100-200 units/kg BD (goal anti-Xa level 0.5-1unit/ml); low dose defined as dalteparin 50 units/kg BD (anti-Xa levels unmonitored)
ECMO: extracorporeal membrane oxygenation; PIMS-TS: paediatric inflammatory multisystem syndrome temporally associated with SARS-CoV-2; SARS-CoV-2: severe acute respiratory syndrome coronavirus.

Underlying comorbidities were common in the respiratory group (9/10), 60% demonstrated more than one co-morbidity. In the incidental group, all but one had an underlying immune-compromising condition (malignancy, metabolic disease, sickle cell anaemia). The remainder of the immune-compromised patients generally had mild disease phenotypes. Conversely, PIMSTS cases were largely healthy at baseline (61% no comorbidities) with obesity making up the majority of the comorbid conditions. Full characterisation of co-morbid states per SARS-CoV-2 disease phenotype are presented in *Supplementary Table A*.

At discharge, 73% of patients were functionally back to baseline, whereas 18% were functionally independent but not back to pre-admission status and/or had minor organ sequelae requiring ongoing follow-up but no active management. Seven percent of patients had major sequelae requiring assistance with activities of daily living and/or requiring active management of end organ damage. One patient with serious complications of PIMS-TS remained an inpatient. No directly-related COVID-19 deaths have been reported.

### (ii) Intensive Care Outcomes

A total of 259 children were admitted to our COVID-19 isolation and non-isolation PICUs from the 26^th^ March to the 19^st^ May 2020. Of all admitted patients to both PICUs, 86 (33%) were suspected to have COVID-19.

36 patients in our cohort required PICU admission (63%) with a median length of stay of three days (IQR 1–7). 21 required mechanical ventilation (37%) with a median duration of two days (IQR 1–6) and a maximum duration of 16 days. The median duration of ventilation for those meeting pARDS criteria (n = 4) was 13 days (IQR). PICU management of hypoxic respiratory failure included: inhaled pulmonary vasodilators, prone ventilation, and high frequency oscillation. Twenty-three patients in our cohort (40%) required inotropes for a median duration of two days (IQR 1–3) with the longest duration of inotropic support reaching 13 days. Inotropic requirement was overwhelmingly more common in the PIMS-TS cohort (65%). One patient with PIMS-TS required extracorporeal membrane oxygenation (VA-ECMO). No patients required dialysis. Of those admitted to the COVID-19 PICU irrespective of final diagnosis, 67% received prophylactic anticoagulation as part of a modification to our normal practice. 81% of PIMS-TS and 60% of respiratory patients were anticoagulated with LMWH, although anticoagulation was not exclusive to the PICU setting. All children survived discharge from PICU.

## Discussion

Our cohort illustrates four distinct SARS-CoV-2 disease groups, with distinct demographic differences who required different management approaches. Dissimilarities in presentation, management, and follow-up of paediatric versus adult cohorts must be considered in both the anticipation of a second wave of COVID-19 and for future pandemic planning.

Compared to adult cohorts, to date, there remains a paucity of data relevant to COVID-19 in paediatric populations even at the conclusion of the initial wave of the pandemic. Notably, there was minimal understanding of paediatric SARS-CoV-2 effects prior to the influx of cases in Europe and America. The largest (n = 2143) and earliest review of paediatric COVID-19 patients in China described a relatively unaffected cohort with 5.6% and 0.6% suffering from hypoxia and multi-organ/pARDS respectively; 4.4% of paediatric patients were completely asymptomatic and 89.7% had only mild/moderate disease (3). Likewise, a Centers for Disease Control and Prevention (CDC) report of 150,000 cases in the United States comprised of only 1.7% paediatric cases, suggesting low incidence in children with less severe pathology (4). Similar rates had been described in Europe with paediatric cases in Italy encompassing only 1.2% of initial COVID-19 patients (5).

Our patient characteristics are similar to a separate UK single centre experience although in this cohort serologic positivity was not an inclusion criterion and children were not grouped by phenotypic presentation (6). Of significance, there was a parallel preponderance for affected children from ethnic minorities. Additionally, a case series of patients admitted to a New York Children’s hospital demonstrated obesity as a similarly prominent risk factor for admission with severe disease, with traditionally at risk paediatric populations relatively spared (7).

As a result of reassuring paediatric data at the start of the pandemic, initial planned patient pathways included transfer of SARS-CoV-2 patients to regional High Consequence Infectious Diseases Units. In order to facilitate expansion of adult inpatient capacity as cases surged in the UK, paediatric secondary care services from the region were relocated to our centre. Limited initial adaptation was made for severe cases of COVID-19 requiring intensive care as, based on the Chinese and Italian data the assumption was that more would not be necessary. However, crucially, protocols were put in place to activate and expand quickly if necessary. At the onset of the unexpected paediatric surge, a separate dedicated 16 bed COVID-19 PICU was created within 48 hours to accommodate increased patient numbers.

Treatment guidelines for all evolving paediatric SARS-CoV-2 phenotypes, based on available evidence, were drafted prior to arrival of the first patients and were given expedited approval by the hospital drug and therapeutics committee. A system for rapid approval of investigational drugs was set up in partnership with the trust bioethics committee with decision-making support from an established multidisciplinary team (MDT) of specialists, inclusive of external specialist support as an impartial presence. In light of the lack of evidence for treatments of this patient group, pooling expertise in the MDT has provided a basis for the development of a standardised treatment protocol upon which an evidenced based approach could be built. The MDT embraced videoconferencing technology to uphold infection-prevention-and-control (IPC) precautions, could be convened urgently by any member ad hoc (sometimes within minutes) to discuss critical cases, or when rapid approval of investigational treatments was necessary.

The complexity of paediatric pandemic preparedness in our centre can be summarised in four distinct ways:

i. *Changing Disease Phenotypes: Evolving presentations with chronologically heterogeneous groupings of childhood disease.* Preparedness for changing pathology required different levels of care, the creation of an MDT, and the construction of separate COVID-19 and non-COVID-19 PICUs with a specific COVID-19 general paediatric step-down ward.
ii. *Unforeseen Disease Pathology: The emergence of PIMS-TS not previously described in the initial Asian epidemic.* Timely collaborative efforts with other paediatric centres capitalising on intra- and inter-institutional multidisciplinary input was essential in the early recognition and management of this novel paediatric phenomenon. As long-term effects remain unknown, the MDT approach will equally be essential in patient follow-up.
iii. *Changing Treatment Modalities: The evolution of COVID-19 treatment evidence extrapolated from adult data at the height of our local epidemic.* There was a timely need to adapt paediatric management pathways in our cohort according to latest research. However, alteration in treatment was carried out cautiously given distinct clinical differences in children versus adults. Moving forward, our cohort re-enforces the need for: paediatric specific clinical trials, inclusion of paediatric patients in large multicentre trials and observational studies, and paediatric-specific pharmacokinetic data for novel drug treatments.
iv. *Changing Patient Demographics: A high proportion of paediatric pathology in adolescent ethnic minority populations.* This must be considered in the development of adequate prevention strategies, tailored medical management, supportive care, and targeted follow-up strategies implemented in a culturally appropriate manner, acknowledging COVID-19 impacts on both physical and mental health.

From a laboratory perspective, rapid upscaling of microbiology and immunology capacity was reliant on the ability to adapt at a fast pace. The on-site laboratory promptly developed validated RT-PCR and antibody testing. Provision of validated serology testing early on in the pandemic, when this was not yet available widely at other centres, was fundamental to the identification of SARS-CoV-2 as the likely trigger for PIMS-TS. In addition, RT-PCR was performed, but yet to be fully validated on: cerebrospinal fluid (CSF), stool, blood, urine, and saliva samples. This has allowed for testing of various body fluids in patients with atypical presentations potentially triggered by SARS-CoV-2. Our immunology laboratory scale-up has also allowed for more widespread and timely assessment of cytokine responses and immunological effects of SARS-CoV-2 in the paediatric population.

## Limitations

Our local demographic population may not be generalisable to other settings. The referral nature of our centre did not allow us to capture data prior to transfer or after discharge back to local hospitals.

## Conclusions

Supported by an ability to adapt quickly in all phases of a pandemic, our COVID-19 paediatric outcomes were reassuringly good. Our centre needed to evolve throughout the SARS-CoV-2 pandemic as new phenotypes arose, those not previously described in other regions, and adjust when new at risk populations were identified. An adaptive, MDT approach was paramount. Expanded laboratory capacity and incorporation of technology platforms to facilitate remote collaboration in response to strict infection control precautions were both indispensable. Paediatric-specific planning must not be static and evolution of preparedness endeavours must continue, particularly in the face of a potential second wave of SARS-CoV-2. Of utmost importance, the distinction between paediatric and adult populations must not be overlooked.

## Data Availability

Anonymised data was collected and stored in a secure Excel database

## Funding Statement

This research received no specific grant from any funding agency in the public, commercial or not-for-profit sectors.

## Contributorship Statement

NA, JP, and KM conceptualised the study. NA, JP, KG, CP extracted patient data. JH extracted neurological patient outcomes. NA performed the statistical analysis. JP and NA were the main authors of the manuscript with considerable contributions from NA, AB, CP, KM, and MJ. All authors were involved in the data collection, drafting, and revision process.

## Acknowledgements

The clinical care provided by all junior doctors, nurses, allied health professionals, health care assistants, and technicians at GOSH was essential in this a novel pandemic. Important members of the MDT included but not limited to: Drs Filip Kucera, Delane Shingadia, Louis Grandjean, Muthana Al Obaidi, Paul Brogan, Charalampia Papadopoulou, Elena Moratis, Sandrine Lacassagne, Imke Meyer-Parsonson, Lee Hudson, Noelle Enright, and Premala Muthukumarasamy. Kimberly Gilmour, Katherine Harris, the GOSH microbiology service, and all laboratory personnel were integral in the development of validated laboratory tests and expansion of laboratory capacity.

## Notes

Conflicts of Interest: AB received funds for consultative work provided to Gilead Sciences Inc. This consultative work did not impact on the results of the manuscript submitted. No conflicts of interest declared.

### Competing Interest Statement

AB received funds for consultative work provided to Gilead Sciences Inc. This consultative work did not impact on the results of the manuscript submitted. No conflicts of interest declared.

### Clinical Trial

The project was registered with the local research department (approval #2857)

### Funding Statement

No funding

### Author Declarations

The project was registered with the local research department (approval #2857)

